# Reorganization of nurse scheduling reduces the risk of healthcare associated infections

**DOI:** 10.1101/19007724

**Authors:** Eugenio Valdano, Chiara Poletto, Pierre-Yves Boëlle, Vittoria Colizza

## Abstract

**Background:** Efficient prevention and control of healthcare associated infections (HAIs) is still an open problem.

**Objective:** To design efficient hospital infection control strategies by reorganizing nurse scheduling.

**Design, setting, and participants:** Proof-of-concept modeling study based on high-resolution contact data from wearable sensors between patients, nurses, doctors, and administrative staff at a short-stay geriatric ward of a University hospital.

**Methods:** We considered isolation and contact removal to identify the most important class of individuals for HAI dissemination. We introduced a novel intervention based on the reorganization of nurse scheduling. This strategy switches and reassigns nurses’ tasks through the optimization of shift timelines, while respecting feasibility constraints and satisfying patient-care requirements. We evaluated the impact of interventions through a Susceptible-Colonized-Susceptible transmission model on the empirical and reorganized contacts.

**Results:** Isolation and contact removal produced the largest risk reduction when acting on nurses. Reorganizing their schedules reduced HAI risk by 27% (95% confidence interval [24,29]%) while preserving the timeliness, number, and duration of contacts. More than 30% nurse-nurse contacts should be avoided to achieve an equivalent reduction through simple contact removal. No overall change in the number of nurses per patient resulted from the intervention.

**Conclusions:** Reorganization of nurse scheduling offers an alternative change of practice that substantially limits HAI risk in the ward while ensuring the timeliness and quality of healthcare services. This calls for including optimization of nurse scheduling practices in programs for better infection control in hospitals.

---

Healthcare associated infections (HAIs) are increasingly widespread, with an estimated 4 million individuals affected each year in Europe, representing approximately 6% of all hospitalized patients (1). These infections have a substantial impact on morbidity and associated costs for the healthcare system, potentially leading to failure of treatment, longer illnesses and hospitalizations, and deaths. Rising antimicrobial resistance in hospitals has also increased the threat to human health, as resistant pathogens may cause serious infections that cannot be treated with available drugs (2).

Common HAIs spread through close-range proximity or physical contacts between individuals. Several studies highlighted the importance of contacts for HAI diffusion (3–9), showing how larger variety and duration of contacts are associated to an increase in HAI risk (10,11). These factors lead to the well-known paradox that healthcare workers play a key role in pathogen dissemination because of their frequent and persistent contacts with individuals of different categories (12). Being at higher risk for HAI colonization, healthcare workers may act as transient superspreaders and transmit the infection to the large number of individuals they get in contact with, especially in the vulnerable population of patients (12–14).

Infection control strategies targeting healthcare workers require careful design, to avoid interfering with their ability to carry out their core healthcare responsibilities. Hygienic measures such as hand sanitizing are the primary strategy to prevent HAI diffusion, aiming to reduce the per-contact risk of transmission (15). The efficacy of these measures is however limited by low compliance rates, as reported by several studies especially under conditions of emergency or understaffing (12,16–18). Even low compliance by a few individuals can have a disproportionate impact on the risk of HAI diffusion in the hospital, given the presence of potential superspreaders (11,12,19,20). Other approaches for infection control have therefore considered the use of personal protective equipment (e.g. face masks and gloves) (21), vaccination (22), isolation, or nurse cohorting (i.e. assigning nurses to a limited number of patients during a given working period) (23). Their effectiveness, however, is still matter of debate (23,24). Most importantly, some of these measures may only be applicable in reaction to outbreaks, as they are rather costly and disruptive. It may thus prove difficult to integrate them into day-to-day hospital activities.

Routine operations in a hospital are ensured by adequate healthcare workers staffing and scheduling. Their organization has been extensively studied for several decades in operations research, management, and computer science (25,26) and is generally known as the ‘nurse scheduling problem’. It typically involves the optimization of single or multiple goals while satisfying a set of hard constraints – i.e. features that need to be respected at all costs, e.g. feasibility, workload, length of shifts, required personnel or skills – and a set of soft constraints – i.e. aspects that are desirable but may not be met in order to achieve a solution, e.g. preferences for a day off. Mathematically described by the constrained minimization of a potential function, the solution to the scheduling problem aims to optimize human resources’ efficiency, patient safety, quality of medical services, costs, and staff satisfaction. Despite the great interest in the topic, research has so far addressed it exclusively from the management and computational perspectives (25,26), with no regards to its potential role in infection control.

Here we propose a proof-of-concept modeling study for hospital infection control based on the reorganization of care in a hospital ward through changes in the schedule of work shifts of nurses. Using high-resolution temporal records on contacts in a hospital ward (3), our approach switches tasks between nurses by altering their work schedules through the optimization of a potential function, similarly to models for nurse scheduling. The reorganized schedule maintains full staff capacity at any given time, preserves all time-referenced contacts recorded in the dataset without affecting quality standards of medical services, and respects basic occupational constraints (weekly workload, length of a work shift). The study is applied to a short-stay geriatric hospital ward in Lyon, France, where contact data were collected through automated sensors (3). We model the circulation dynamics of hand-transmitted pathogens such as methicillin-resistant *Staphylococcus aureus* (MRSA) or vancomycin-resistant *Enterococci* (VRE) in the ward, and evaluate the effectiveness of the intervention by measuring the risk for HAI diffusion.

## METHODS

### Contact data

We used publicly available anonymized data collected during 4 days and 4 nights (December 6 to 10, 2010) at a short-stay geriatric ward of a hospital in Lyon, France (3,27). Using wearable RFID sensors, the system tracked face-to-face proximity contacts over time between 75 participating individuals, including 27 nurses (N), 11 doctors (D), 8 administrative staff (A), and 29 patients (P). The dataset was first analyzed in Ref. (3); Figure 1 reports its basic properties. Nurses and doctors had the largest cumulative duration of contacts, and most frequent contacts were observed between nurses (NN), and between patients and nurses (PN).

**Figure 1:**
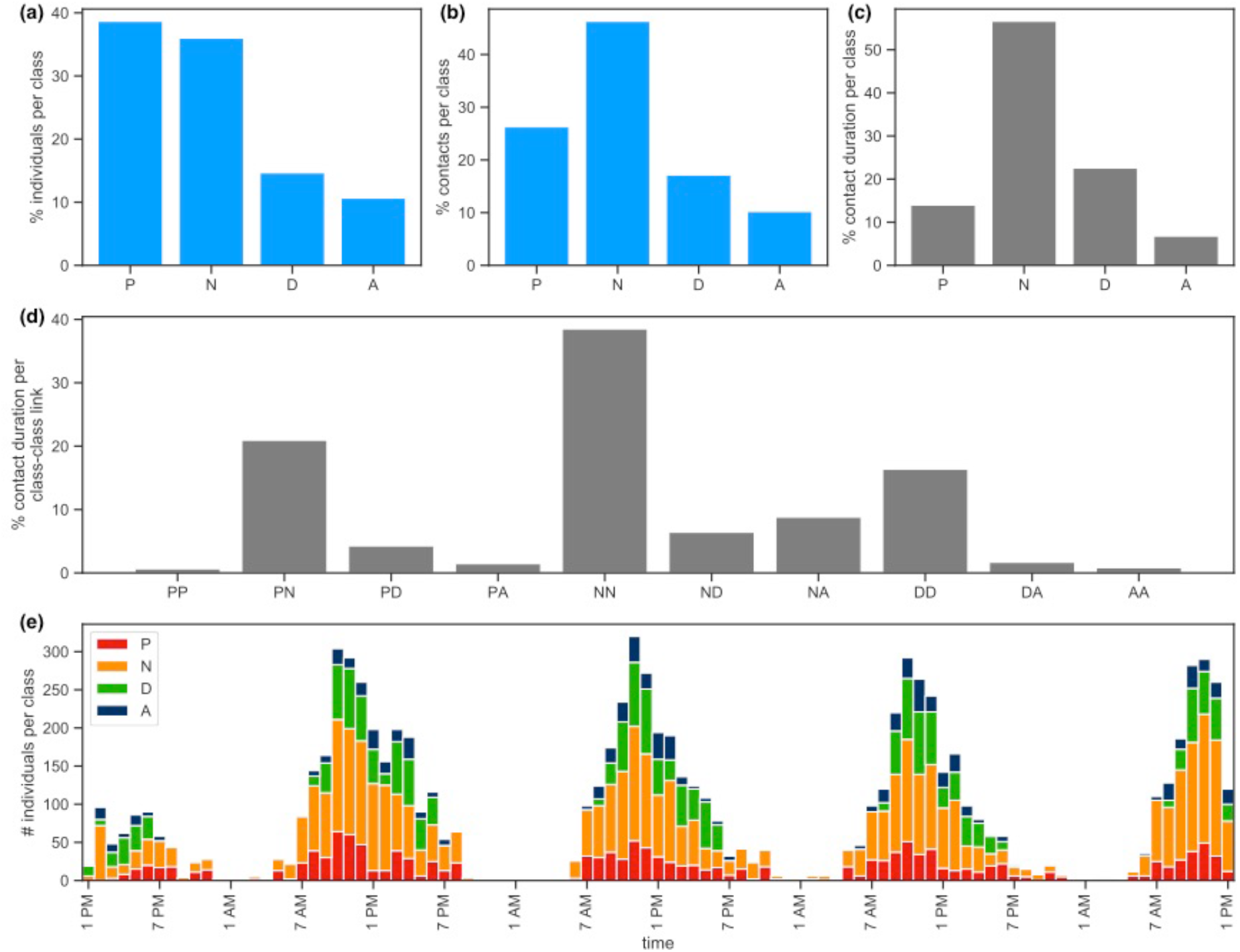
Contacts in the hospital ward. Percentage of participating individuals (a), of contacts (b), and of contact duration (c) by class of individuals (patients (P), nurses (N), doctors (D), administrative personnel (A)). (d): Percentage of contact duration between classes of individuals. (e): Hourly timeline of the number of individuals per class establishing contacts.

### HAI risk estimate

The time-resolved contacts are represented in the form of a temporal contact network (28), where nodes correspond to individuals and links to proximity encounters. Time evolution occurs at an hourly timescale. We model HAI diffusion in the hospital ward through a Susceptible – Colonized – Susceptible transmission dynamics on the temporal contact network (6,8,19,29,30). Colonized individuals transmit the pathogen with probability λper contact. Their average colonization duration is fixed at 10 hours for healthcare workers, assuming a spontaneously clearing transient colonization at the end of a work shift. The duration is longer for patients and corresponds to 10 days, hypothesizing a weekly bacterial screening, followed by 3 days to obtain test results and implement a decolonization therapy, as in (6,19).

To assess the risk of transmission of the infection in the ward, we estimate the condition for circulation of MRSA or VRE on measured contacts through the infection propagator approach (31–33). This theoretical framework was introduced to study epidemics spreading on temporal networks and identify the critical value λ_*c*_of the transmissibility above which the pathogen spreads in the host population (i.e. if λ>λ_*c*_an outbreak is predicted to occur). The Appendix reports a full description of this approach, and the available software tool.

### Intervention through isolation or contact removal

To assess the role that each class of individuals has on HAI risk, we simulate two interventions based (i) on the isolation of individuals belonging to a given class, and (ii) on the removal of contacts established between two classes (e.g. contacts between patients and nurses). Each intervention is made comparable across classes or pairs of classes, through the isolation of 8 individuals (i.e. the smallest size class) or the removal of 5% of the total duration of contacts in the dataset, respectively. Interventions are repeated to account for the stochasticity in the choice of the node to isolate or of the contacts to remove.

### Intervention based on reorganization of nurse scheduling

We introduce an activity variable *a*_*i*_(*t*) associated to nurse *i*during hour *t*, so that *a*_*i*_(*t*)=1 if the nurse is at work and establishes contacts in that hour and *a*_*i*_(*t*) =0otherwise (Figure 2a). For each nurse, we compute the shift duration *s* defined as the number of consecutive work hours, and the workload *w*corresponding to the total number of hours worked in the dataset.

**Figure 2:**
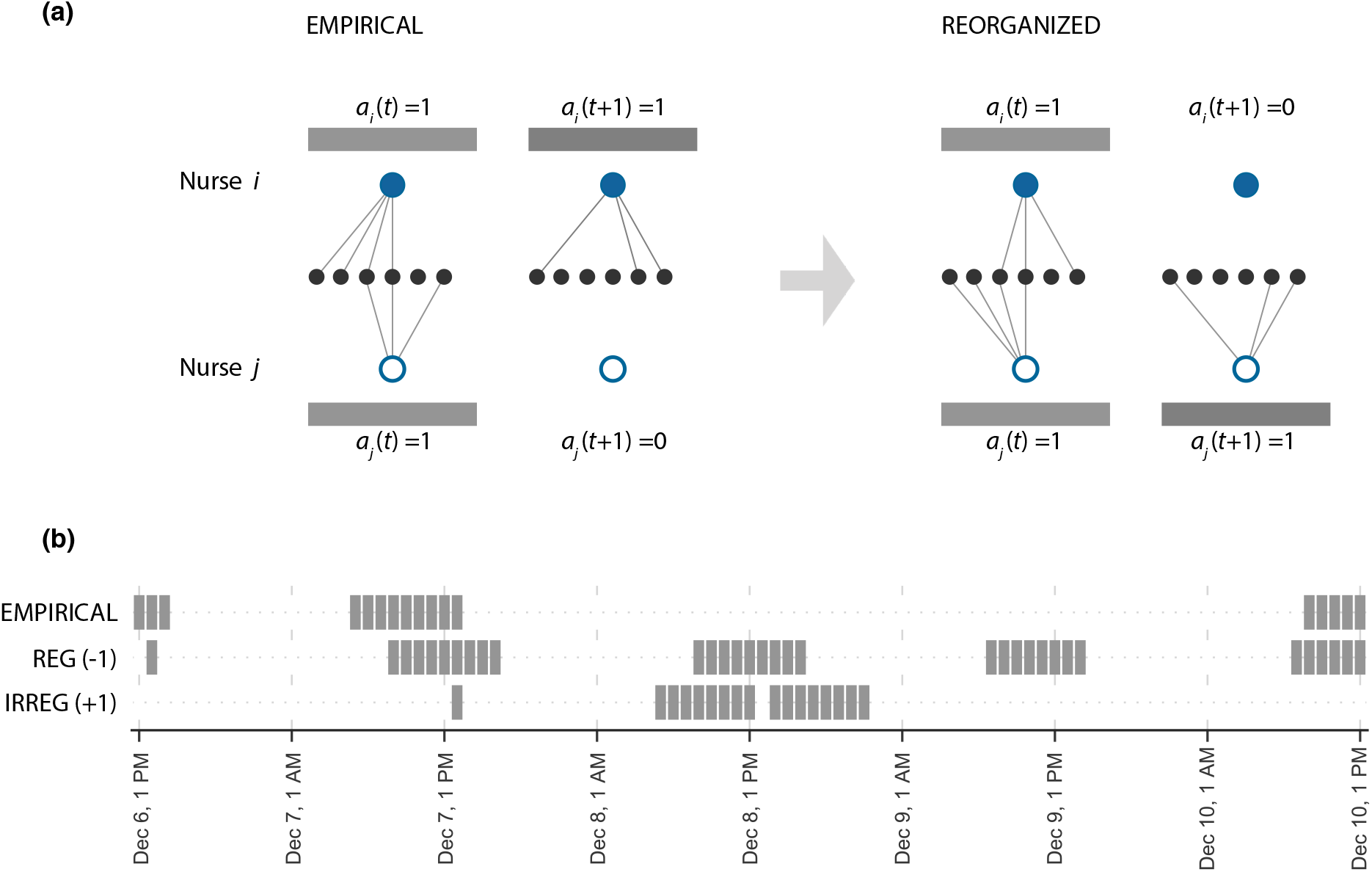
Intervention on nurse scheduling. (a) Schematic visualization of the intervention, with the exchange of tasks between nurse *i* (filled blue node) and nurse *j* (void blue node), at times *t* (while they are both at work) and *t* + 1(while nurse *j* is not at work in the empirical schedule, and would exchange her shift with nurse *i* in the reorganized schedule). Links represent contacts with other individuals (black nodes). The reorganized nurse schedule (right) is compared to the empirical one (left) obtained from the contact data. (b) Example of an empirical nurse schedule along with the re-organized ones obtained with *k*=−1and *k*=−1, leading to regular and irregular individual schedules, respectively. Grey blocks correspond to hours when the nurse is at work.

The proposed intervention switches and reassigns the tasks performed by two nurses in a given hour *t*. Tasks consist in contacts that nurses establish, as they perform their duties in interaction with other individuals (e.g. caring for a patient). They correspond to possible transmission events. The reorganization is driven by the minimization of the following potential function:

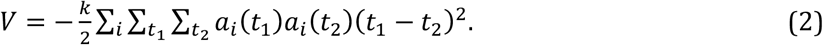

*i* runs on all nurses, *t*_1_and *t*_2_run on the whole timeline, and *k* determines the tendency of the potential (*k* = ±1). *k* =−1shows a tendency to create *regular* individual schedules (periodic activity patterns), and *k*= 1a tendency for *irregular* individual schedules (erratic activity patterns) (Fig.2b).

This strategy preserves the number, type, and exact timeline of contacts, differently from the intervention through contact removals. The minimization of the potential is additionally subject to feasibility constraints on shift duration and workload of nurses:

- Model *S*: the exchange is allowed as long as each working shift lasts at most *s=*10 hours, as measured empirically.
- Model *WS*: in addition to the constraint on shift duration, the exchange is allowed only if it preserves the empirically measured workload *w* of each nurse.

Each model is run with both values of *k*, for a total of 4 reorganization options (*S*_-1_,*S*_+1_,*WS*_-1_,*WS*_+1_). The Appendix reports a detailed description of the minimization algorithms. Despite being synthetic, these interventions have an increasing degree of realism to show the potential of this proof-of-concept study for possible applications in real situations.

### Evaluation of interventions

We evaluate the effect of interventions by comparing the resulting HAI risk estimate 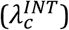 with the one estimated on the empirical pattern of contacts 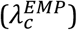. We define the *HAI risk reduction* as the relative variation of these two quantities 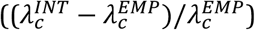, so that a positive risk reduction corresponds to interventions improving the control of potential HAI diffusion in the hospital ward (the opposite for negative values). Fluctuations in the HAI risk reduction are obtained from the variations resulting from the stochastic trials.

### Effects of the reorganization of nurse scheduling on contact patterns

To test whether the proposed reorganization of nurse scheduling leads to nurse cohorting (23), we measure the variation in the number of distinct nurses assigned to each patient following the intervention compared to the empirical value. Negative values of this variation correspond to nurse cohorting (i.e. an average reduction of the number of nurses per patient).

We also measure the variations in the nurses’ degree (i.e. number of distinct connections each nurse establishes) by comparing average degree and associated fluctuations before and after the reorganization.

## RESULTS

### Intervention through isolation or contact removal

Complete isolation of 8 randomly chosen patients corresponds to a drop of 8% in the cumulated duration of contacts, while isolating 8 nurses reduces it by 31% (Figure 3a). In the latter situation, it leads to an approximate 35% median reduction of the HAI risk (Fig. 3c), whereas the same intervention applied to other classes has negligible impact.

**Figure 3:**
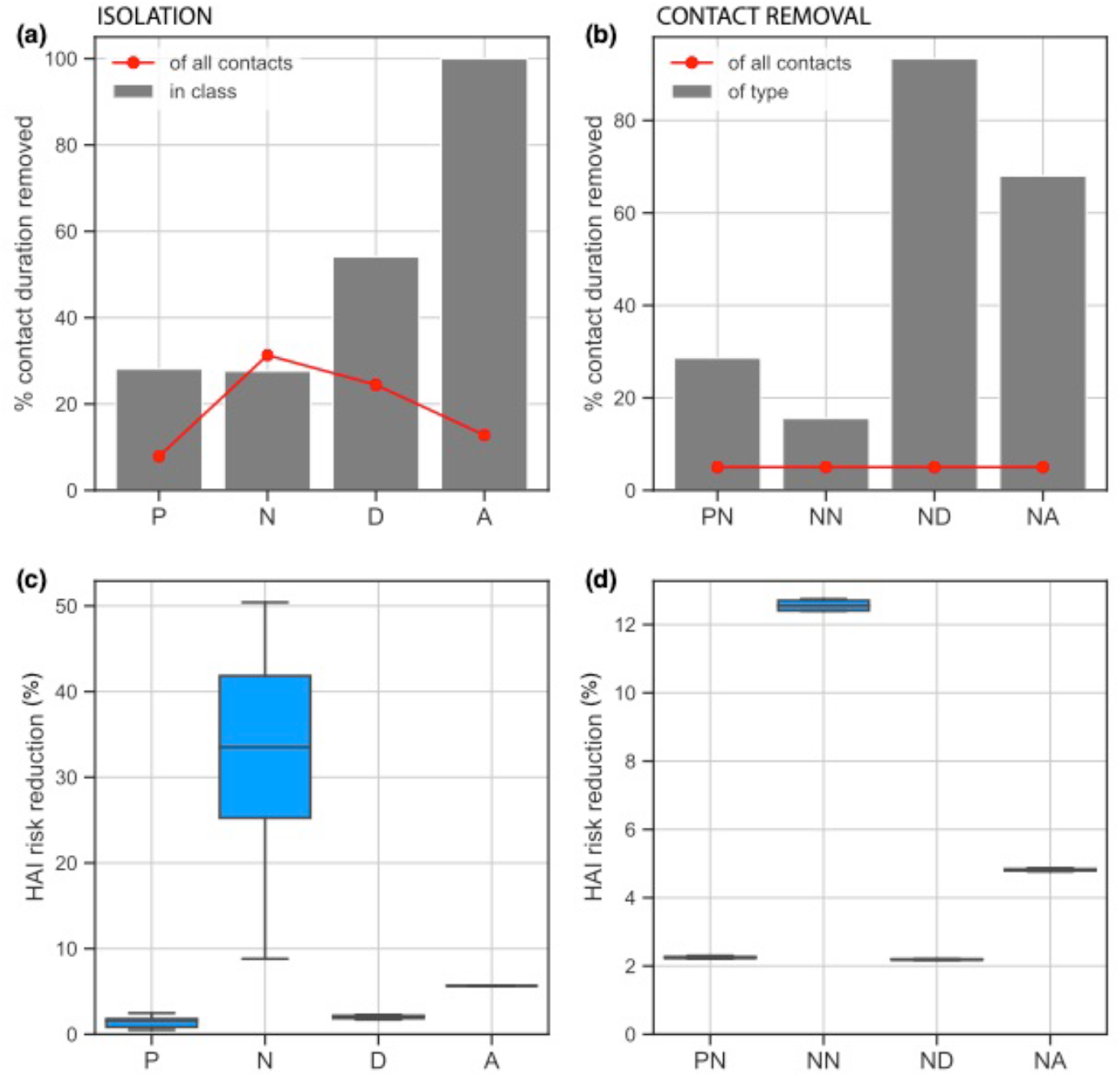
Impact of intervention through isolation or contact removals. (a): Median percentage of contact duration removed within the class (red line) or of the full timeline (grey bars) once 8 individuals in each class are isolated, corresponding to 28% of patients (P), 30% of nurses (N), 73% of doctors (D), and 100% of administrative staff (A). (b): Median percentage of contact duration removed among links established by nurses with other classes (grey bars, with patients (PN), with nurses (NN), with doctors (ND), with administrative staff (NA)), once 5% of the total contact duration of the full timeline is removed (red line). (c), (d): HAI risk reduction in the hospital ward achieved through isolation (panel c) or contact removal (panel d) corresponding to the results of panels a and b, respectively. Boxplots indicate the median, interquartile range and 95% CI of the risk reduction, accounting for the stochasticity of the interventions (results from 20 random trials).

When removing a certain fraction of contacts between classes, the largest risk reduction is obtained by acting on nurse-nurse contacts (median reduction of 13%), corresponding to deleting 15% of nurse-nurse contact duration (Fig. 3b, d). Interventions on contacts between nurses and doctors or administrative staff, which are proportionally more disruptive, have almost no impact on the risk.

Both theoretical interventions highlight the central role played by nurses in the hospital ward under study, supporting the design of a more realistic intervention that could act on nurse activities without disrupting the ward functioning and the provision of medical and nursing services.

### Intervention based on reorganization of nurse scheduling

Minimizing the potential while constraining only the maximum shift duration leads to two different profiles of the workload distribution (Figure 4). Model *S*_-1_shows approximately half of the nurses not working (*w*=0), and the rest distributed quite evenly from short to very long workloads (Fig. 4a). Model *S*_+1_tends instead to homogenize nurses’ workload around the average value (16 to 23 hours in the 4-day timeframe, Fig. 4c).

**Figure 4:**
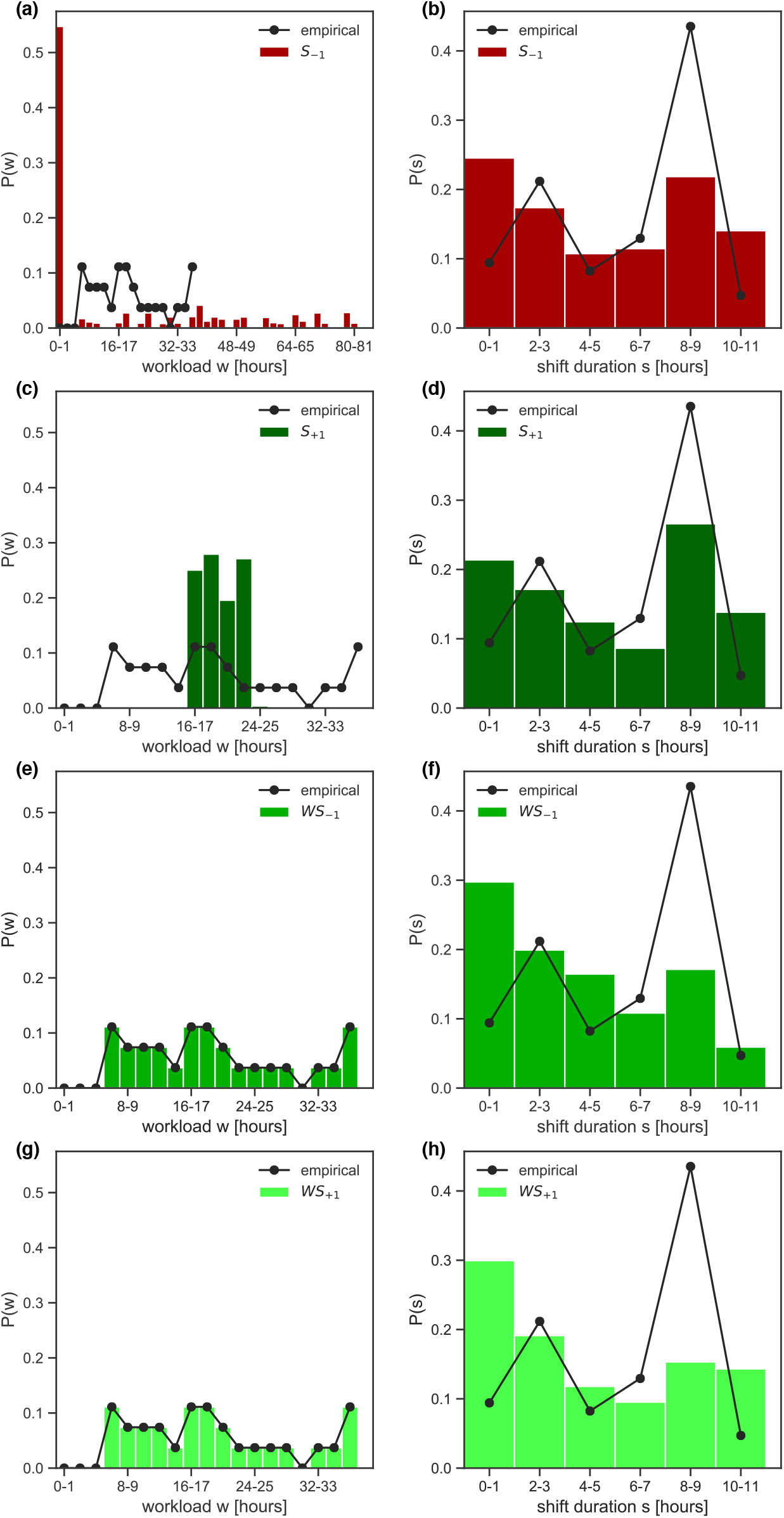
Workload and shift duration in the reorganized schedule. (a)-(b): Probability distribution of nurses’ workload *w*(panel a) and shift duration *s*(panel b) following the reorganization of shifts, compared to the empirical distributions. The reorganization is based on model *S*_-1_(i.e. constraint on shift duration and attractive potential). (c)-(d), (e)-(f), (g)-(h): As panels (b) and (c) for model *S*_+1_(constraint on shift duration with repulsive potential), model *WS*_-1_(constraint on workload and shift duration with attractive potential), model *WS*_+1_ (constraint on workload and shift duration with repulsive potential), respectively.

Shift duration distributions are rather similar in all models, and qualitatively comparable with empirical data (Fig. 4b, d, f, h). Small variations on 1-hour shifts (higher probability in *WS* models) and 8-9-hour shifts (more marked increase in the *S* models) are observed.

Model *S*_+1_achieves the largest reduction of HAI risk (median 27% reduction, 95% CI [24,29]%), followed by *WS*_-1_(21%, [20,24]%) and *WS*_+1_(19%, [16,20]%) (Figure 5a). Equivalent risk reductions would be obtained by contact removal if more than 30%, 25%, and 20% duration of nurse-nurse contacts were to be removed, respectively (Fig. 5b). Model *S*_-1_instead increases HAI risk of 5%.

**Figure 5:**
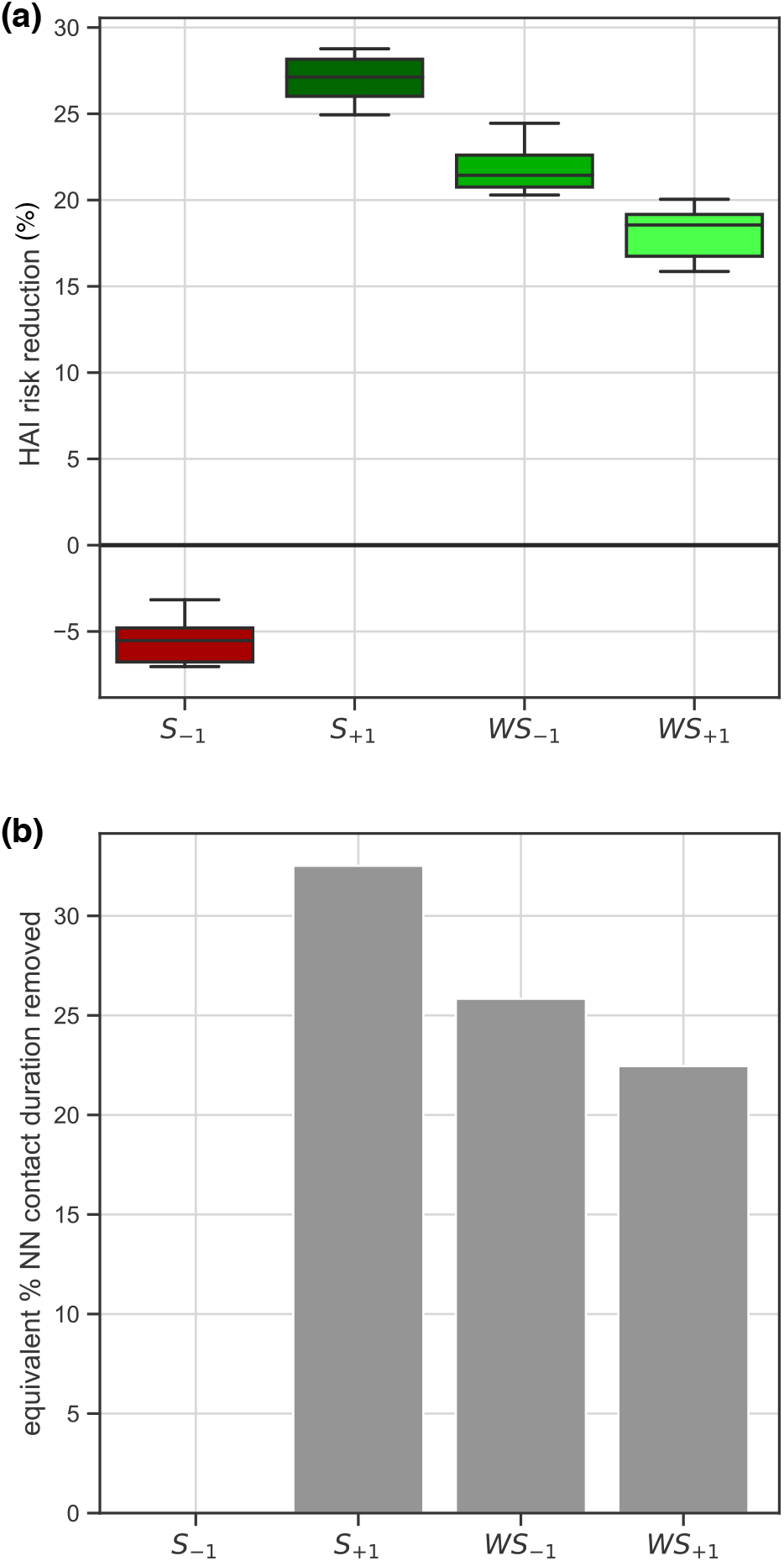
Impact of intervention through reorganization of nurse scheduling. (a): HAI risk reduction in the hospital ward achieved with the reorganization of nurse scheduling in the models *S*_-1_,*S*_+1_,*WS*_-1_,*WS*_+1_, compared to the empirical situation. Boxplots indicate the median, interquartile range and 95% CI of the risk reduction, accounting for the stochasticity of the exchange (results from 50 random trials). (b): Percentage of contact duration to be removed in the nurse-nurse interactions so that the intervention through contact removal would achieve the same risk reductions of panel a (obtained through the reorganization of nurse scheduling). *S*_-1_is not shown as it has a negative impact on the risk.

Models *S*_+1_, *WS*_-1_, and *WS*_+1_, which decrease risk, show a reduction of the fluctuations in the number of distinct contacts established by nurses, without substantially altering their average number of contacts or the number of distinct nurses assigned to each patient (Figure 6). Model *S*_-1_, which increases risk, raises cohorting levels with a median of 4 less nurses assigned to each patient and strongly increases nurses’ degree fluctuations.

**Figure 6:**
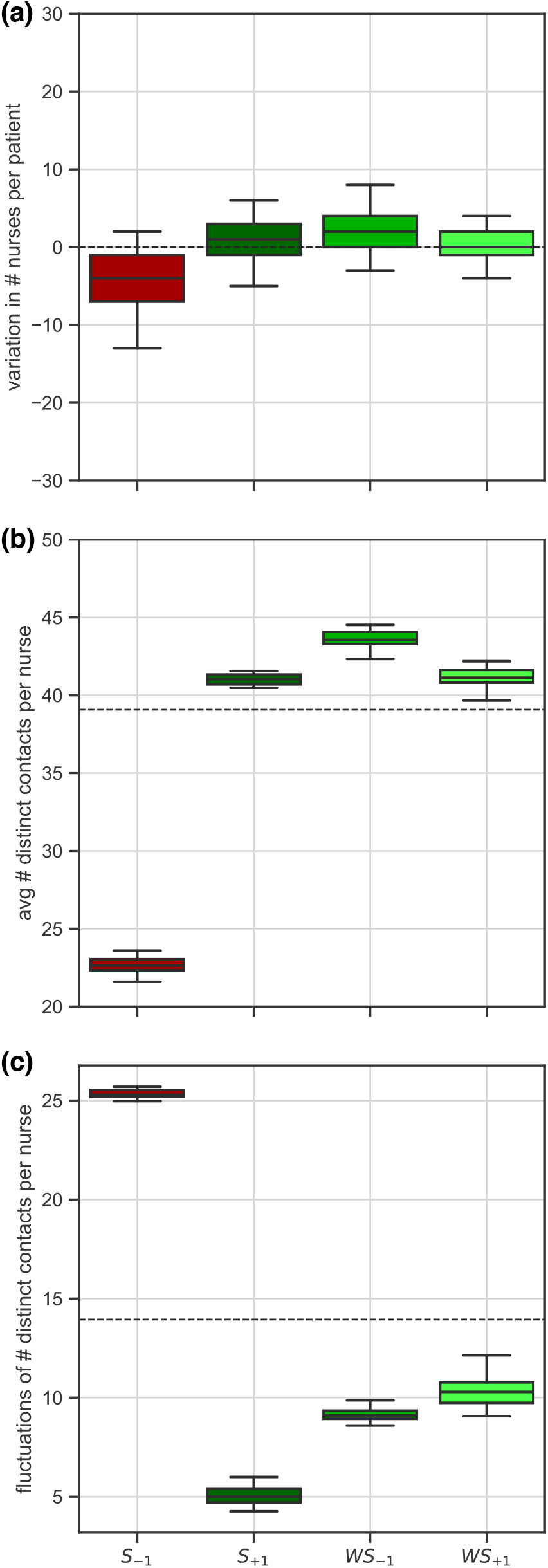
Effect of reorganization on contact patterns. (a) Variation of the number of distinct nurses assigned to each patient, in the reorganized vs. empirical contact pattern. (b) Average number of distinct contacts per nurse (nurses’ degree). Dashed line corresponds to the empirical value. (c) Fluctuations (standard deviation) of the number of distinct contacts per nurse. Dashed line corresponds to the empirical value. For (a),(b),(c), results are obtained from 500 random trials.

## DISCUSSION

The key role of healthcare workers in the transmission of healthcare associated infections is widely recognized (12–15). Low compliance and limited sustainability of recommended strategies hinder efficient infection control. Our study proposes an alternative change of practice through the reorganization of nurse work shifts to reduce HAI risk. Using sensed contact data in a hospital ward, we show that reassigning tasks to nurses minimizing a potential function on their timeline of activity can reduce the risk of HAI diffusion by about one third. Our findings show the potential of planning nurse schedules to improve infection prevention and control.

The key advantage of the proposed intervention is that it preserves the number, type, and duration of contacts at each time. This ensures the timeliness and quality of medical and nursing services provided. An equivalent impact on risk reduction could be achieved by limiting the interactions viable for transmission. Uninformed removal of contacts would be however rather disruptive, with one third of contacts deleted among nurses, at the expense of standards of care. Patient isolation, staff cohorting, and increase in staff levels were shown in previous work to be effective in limiting transmission, by directly or indirectly acting on the interactions (18,23,29). Fully isolating about 30% of patients in the ward, however, had no impact on the risk of transmission in this study. Also, the highest improvement in infection control (*S*_+1_) was due neither to cohorting nor to an increase in staff levels. However, this intervention led to the strongest reduction in the fluctuations in the number of distinct contacts per nurse.

Homogenizing nurses’ contact patterns around 40-45 contacts per nurse removes the presence of potential superspreaders (11,12,19,20) that could otherwise act as risk amplifiers. The reduction in the degree fluctuations is indeed observed only in the models reducing HAI risk.

Adequate staffing levels and reasonable workloads are established factors promoting infection prevention in hospitals (12,18,34). Reorganizing nurses’ shifts just respecting the maximum shift duration constraint (model *S*_-1_) results in our study in approximately half of the staff not working, while nursing care is assigned to the remaining half, thus forcing unrealistic work schedules (up to 80 hours of work per nurse in 4 days, i.e. an average of 20h/day). Such reorganization of work has a negative impact on infection control, in line with empirical findings that recognize high workloads, understaffing, and the presence of superspreaders as key risk factors for MRSA circulation in healthcare settings (12,34). In addition, poor infection control would be here associated to an increase of cohorting levels, which simply results from lower staffing. All other models lead instead to an important reduction of HAI risk, with the reorganization being able to break potential chains of transmission through the swapping of tasks. Improved control is achieved by reducing the presence of superspreaders in the ward, under both regular and irregular individual work timelines of nurses, and with different shift and workload distributions. These findings uncover the practical mechanism for improved control and highlights the robustness of the proposed strategy to different requirements on the organization of the workforce.

Mathematical models have already been used to improve our understanding of hospital epidemiology (35,36). They are nowadays increasingly data-driven thanks to remote sensing, allowing an automated collection of close-proximity interactions between individuals, not affected by reporting or observer biases inherent to other approaches (37). This type of contacts was recently shown to explain the diffusion path of several HAIs (6,7,10,38). For this reason, our findings extend to pathogens other than MRSA and VRE, spreading along the same routes, under the hypothesis of relatively rapid decolonization (6,19). Durations of the order of months that are empirically observed in absence of interventions (39) were not examined here because considered inappropriate in the hypothesis of decontamination taking place.

Prior modeling work generally relied on numerical simulations of HAI spread (35). We used the infection propagator approach to estimate HAI risk reduction in a reliable and computationally fast way. This approach was already used to estimate the risk of disease persistence in other epidemic contexts (32,33), and has the advantage of being flexible to the integration of heterogeneities in the force of infection that may depend, for example, on class-specific transmissibility.

The proposed reorganization of staff schedules focused on the class of nurses, as theoretical results on isolation or contact removal clearly identified nurses as the category of healthcare workers who mostly contribute to the transmission risk in the ward. This can be traced back to the larger number and longer duration of contacts established by nurses, commonly required by nursing care, and it is in line with previous observations and modeling works proposing nurses as target group for prevention measures (7,9,22).

Our findings show the potential to integrate infection prevention into the nurse scheduling problem, originally designed to optimize hospital workforce. However, some steps are still needed to carry this novel paradigm into practice, in the form of a roadmap to a future hospital protocol. First, contact data collection on a longer timeframe is required to provide a comprehensive measurement of the functioning of the healthcare setting under study. Moreover, collecting metadata on the type of clinical interventions performed in the ward, the specific roles of subclasses of personnel (e.g. nurse types), the type of patients admitted, and the standard and organization of care (e.g. scheduling practices along the 24 hours) is key to improve the parameterization of the epidemic transmission model and define the conditions for task’ reassignments (e.g. by swapping similar tasks, or tasks that can be handled by the same staff type). Most importantly, such additional data will help constrain the potential function to patient needs and staff requirements. We list, for example, the length of shifts, the number of weekends worked, the number of on/off days, the role of additional personnel (e.g. physical therapists, nutritionists, with different working patterns), the use of part-time and temporary nursing personnel (thus introducing staff for substitute shifts). Integrating these elements would make the re-scheduling feasible, without altering the core of the strategy proposed here.

We presented modeling evidence that reorganizing nurse scheduling while maintaining the number, timeliness, and quality of medical services provided by nursing staff can strongly decrease the risk for HAI diffusion in the hospital ward. Our study provides the theoretical basis for a new control paradigm, showing its potential for integration in future nurse scheduling practices for the implementation of successful infection control programs at the hospitals.

## Data Availability

Data have been made publically available by data owner

## Thank you notes

We acknowledge the Scientific Evolutionary Writing workshop (www.sew-workshop.org) where part of this paper was written.

## ETHICS STATEMENT

The contact data used in this study were collected in a research approved by the French national bodies responsible for ethics and privacy, the “Commission Nationale de l’Informatique et des Libertés” (CNIL, http://www.cnil.fr) and the “Comité de Protection des personnes” (http://www.cppsudest2.com/) of the hospital.

### APPENDIX

#### 1. Infection propagator approach to evaluate the HAI risk

We represent the time-evolution of contacts in terms of a temporal network with adjacency matrices *A*_*t*_, with *t* running on the 1-hour time steps. The entry *i,j*of *A*_*t*_ is equal to one if nodes *i,j*establish a contact during time step *t*, zero otherwise. We model the spread of the pathogen using a Susceptible – Colonized – Susceptible model. A colonized node in the network transmits the pathogen to a connected node with probability λ(transmissibility) at each time step. It also clears the pathogen with a probability *μ*at each timestep. *μ*^-1^is then the average carriage period. There exists a critical value of transmissibility λ_*c*_– called epidemic threshold – that determines the global behavior of the outbreak. If transmissibility is higher than the epidemic threshold (λ>λ_*C*_), introducing the pathogen into the hospital ward is likely to cause a large-scale outbreak. Instead, if transmissibility is lower than the epidemic threshold (λ<λ_*C*_), the outbreak is likely to affect few individuals. Therefore, computing changes in the epidemic threshold is a a synthetic and easy-to-interpret way to weigh the impact of any policy, on the vulnerability of the ward to the pathogen considered. If the epidemic threshold increases following intervention, the ward becomes more resilient to pathogen introduction. Oppositely, if the epidemic threshold goes down, the ward becomes more prone to large-scale outbreaks. This is the rationale behind our definition of HAI risk reduction: 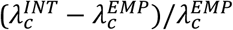. In order to compute it, we need to compute the epidemic threshold before and after intervention. To that end, we employ the infection propagator approach (31–33), which can compute the epidemic threshold on any arbitrary temporal network, for the spreading model used here. The infection propagator is the following matrix:

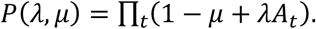

It contains both the time-evolving structure of the contact network (*A*_*t*_), and the parameters of the spreading model (λ,*μ*), and measures the chains of infection between individuals along which the pathogen can spread. We prove in (31–33) that the epidemic threshold is the smallest value of λfor which the largest eigenvalue of *P*equals one.

We provide a Python library to compute the epidemic threshold of any empirical temporal network in the following repository: https://github.com/eugenio-valdano/threshold

#### 2. Implementation of switch and reassignment of nurses’ tasks and minimization of the potential

We minimize the potential using the Metropolis algorithm. It is an iterative process based on the following steps.

Version for *S*_+1_,*S*_-1_:

1. Choose two nurses (*i*≠*j*), and one time step (*t*);
2. If neither nurse is active during *t*, go to 1);
3. Swap tasks between *i,j* during *t*;
4. If the swap breaks the *S* constraint, go to 1);
5. Compute the potential;
6. If the swap decreases the potential, accept the swap. If the swap increases the potential, accept it with probability *e*^-Δ*V*^, where Δ*V*is the change in potential due to the swap;
7. Only if the swap is accepted, update nurses’ task assignments, and potential;
8. Go to 1).

Version for W*S*_+1_,*WS*_-1_:

1. Choose two nurses (*i*≠*j*);
2. Choose two time steps (*t,s*), so that *i*is active during *t*, and not active during *s*, and *j* is active during *s*, and not active during *t*. If this is not possible, go to 1);
3. Swap tasks between *i,j*during both *t,s*;
4. If the swap breaks the *S* constraint, go to 1);
5. Compute the potential;
6. If the swap decreases the potential, accept the swap. If the swap increases the potential, accept it with probability *e*^-Δ*V*^, where Δ*V* is the change in potential due to the swap;
7. Only if the swap is accepted, update nurses’ task assignments, and potential;
8. Go to 1).

